# High Speed Photographic Detection of Particle Distribution During Aerosol and Smoke Generating Surgical Procedures: Effects of Surgical Site Evacuation

**DOI:** 10.1101/2021.05.31.21258042

**Authors:** Kody G. Bolk, Michael E. Dunham, Kevin F. Hoffseth, Jangwook P Jung, Beatriz M. Garcia, Rohan R. Walvekar

## Abstract

**Objective:** Determine the effectiveness of evacuation systems designed to clear bioaerosols and smoke from the surgical field.

**Study design:** High-speed photographic evaluation of aerosol and smoke generated in simulated surgical fields.

**Materials and methods:** Surgical site aerosol clearance was evaluated using a model of the anterior neck and prototypes for surgical site evacuator ports created using 3D printing. A commercially available electrocautery handpiece fitted with an evacuator was tested on animal tissue for smoke clearance. Both systems were connected to a commercial vacuum powered evacuation system. High speed photography was used to record videos of the aerosols and plumes. Fields were recorded with and without evacuation.

**Results:** Efficient aerosol clearance from an open surgical field using an evacuator port is dependent upon the port design, airflow velocity, and placement relative to the aerosol generating site. The size and surface geometry of the surgical field are also important.

Surgical smoke generated with electrocautery is cleared from the field by the evacuation enclosure around the handpiece, even at high electrocautery power settings. Except for device noise, there appears to be no reason for using evacuator flow rates below the maximum setting.

**Conclusions:** Bioaerosol and smoke generated during surgery are potential sources of respiratory pathogens and pose a threat to operating room personnel. Surgical site evacuation can significantly reduce the volume of airborne particles in the field but requires careful design and deployment considerations.x

## Introduction

The COVID-19 pandemic has increased the awareness of pathogenic bioaerosols during aerosol generating procedures (AGPs). The highly contagious SARS-CoV-2 virus is transmitted primarily through respiratory secretions^1^ and is concentrated in the nose, nasopharynx, and oral pharynx, even in asymptomatic patients^2,3^. Health care workers in the operating room who perform procedures associated with high concentrations of the virus in aerosols, such as ear, nose, and throat surgery, are particularly at risk for infection.

Surgical smoke generated during surgery with heat-producing instruments has been well documented as a risk to exposed operating room personnel. Sources include electrocautery, radiofrequency ablation and laser devices. Surgical smoke is approximately 95% water with the remaining 5% consisting primarily of hydrocarbons, tissue particles, viruses, and bacteria^4^. Cultures harvested intraoperatively have documented the presence of live Staphylococcus, Corynebacterium, and Neisseria in surgical plume^5^. Potential viral pathogens include HPV and HIV^6^. Toxic / carcinogenic hydrocarbons found in surgical plume include carbon monoxide, benzene, and hydrogen cyanide^7^.

A direct approach to minimizing the risk of aerosol and smoke producing surgical procedures exists – evacuate the particles at the surgical site. In this study we used high speed photography (HSP) to evaluate the effectiveness of a filtered, vacuum powered evacuator to clear potentially harmful aerosols and smoke from the surgical field.

## Materials and Methods

### HSP testing platform (figure 1)

High speed photographic videos were recorded lateral (0 deg) and overhead (90 deg) relative to the plane of the surgical field. The optomechanical equipment to study aerosols and smoke in the surgical field consisted of a high-speed camera (Phantom T-1340, Vision Research) fitted with a 100 mm lens (Zeiss Optical), and high-intensity (7700 lumens) LED continuous lighting (Multi-LED, GSVitec). Negative pressure was applied to the evacuators using a surgical smoke evacuation system (Smoke Shark II, Bovie Medical). For optimal field of view, the distal end of the camera lens was placed 46 cm from the center of the surgical field for the lateral view and 58 cm for the overhead view. The LED light source was placed at a 15-degree angle to the camera axis. Images of aerosol and surgical smoke created in the surgical field were recorded at 2000 frames per second with an exposure index of 1600 and a frame exposure time of 10 msec.

### High-speed photographic evaluation of surgical site evacuation during aerosol generating procedures

The surgical field of a tracheostomy with an open stoma was simulated using a life-sized manikin of the head and neck (figure 2). The anatomical features were 3D printed from a CT scan-generated computer model. An electronic nebulizer (Vios Pro, Pari) was attached to the internal opening of the stoma aperture.

**Figure 1.**
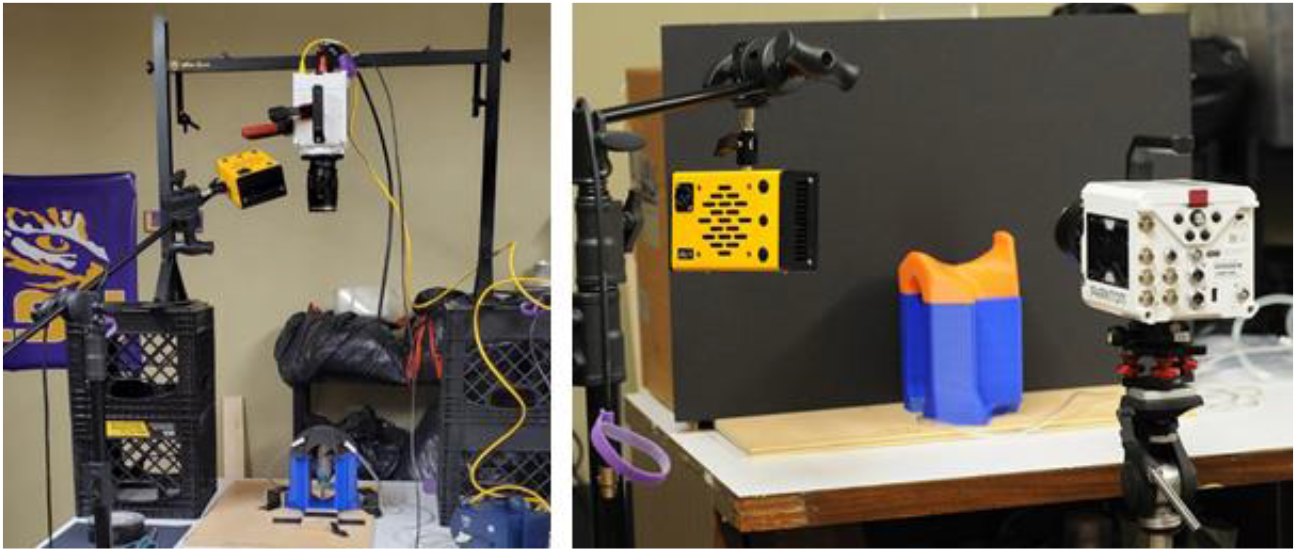
High speed photography rig for recording simulated surgical aerosols and smoke.

**Figure 2.**
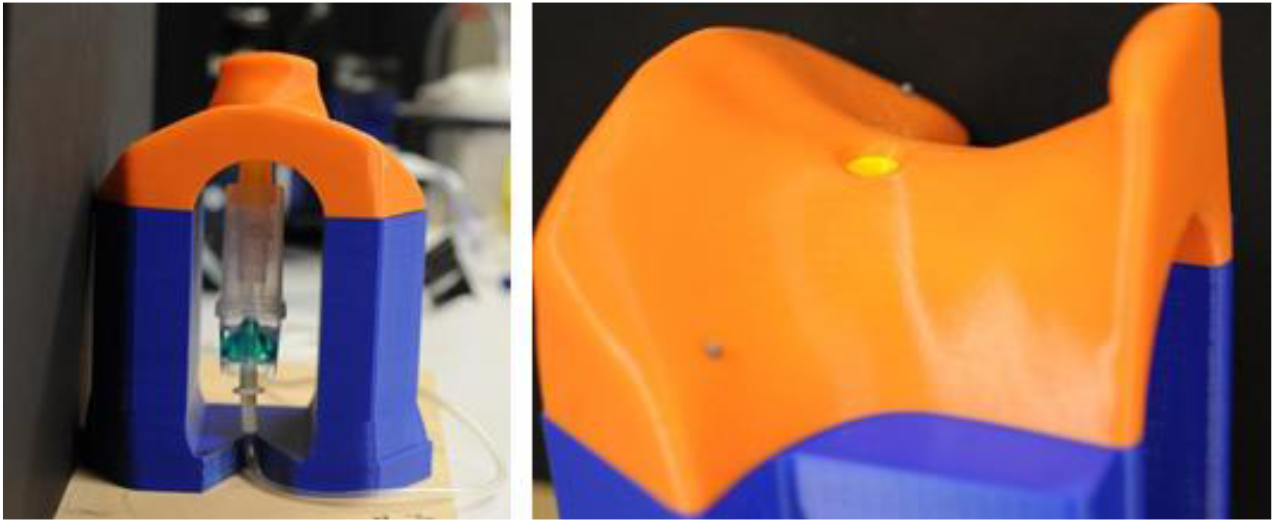
Cervical manikin with nebulizer to simulate stomal aerosols.

The nebulizer chamber was modified to direct 30% of its flow into the cervical stoma. The remaining flow exited the nebulizer from a port on the side of the chamber and out of the open end of the cervical model, consistent with tracheal airflow. The nebulizer console provided pressured air at a flow rate of 6 L/min, resulting in a stomal airflow of 2 L/min. At the stoma (10 mm diameter), this corresponds to a flow velocity of 0.1 m/s. Assuming a tidal volume of 500 ml/breath and a respiratory rate of 12/min, the nebulizer generated flow is consistent with physiologic flows through the tracheal airway during positive pressure ventilation under general anesthesia.

Three port designs were tested (figure 3). The evacuator port was connected to the manikin with adhesive to ensure consistent placement relative to the aerosol. A dual port design was also tested. Port placement reflected the need to avoid interference with surgical access to the field.

**Figure 3.**
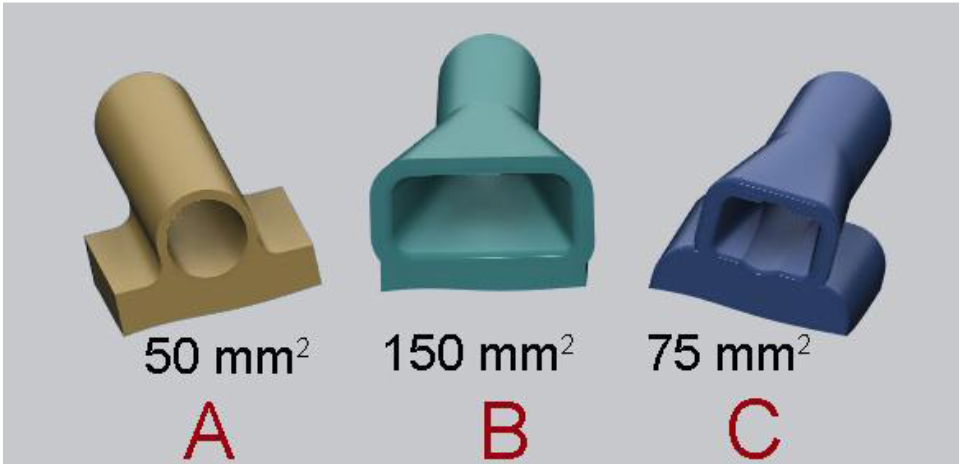
Surgical site evacuator port designs.

The evacuator port was connected to the vacuum console through 9.5 mm or 22 mm corrugated plastic tubing (internal diameter). When the evacuator was set on the maximum setting, the volumetric flow rate was 130 L/min for the 9.5 mm connection and 708 L/min for the 22 mm connection (manufacturer supplied values).

### High-speed photographic evaluation of surgical smoke evacuation created by electrocautery

To study surgical smoke evacuation, we used a commercial electrosurgical handpiece fitted with a smoke evacuation enclosure (Orca, Bovie Medical) (figure 4). The hand piece was connected to its matching electrosurgical console capable of cutting, coagulating and blend frequency settings. The evacuation enclosure was connected to the evacuator through 22.5 mm plastic corrugated tubing. With the evacuator set on the maximum setting, this resulted in volumetric flow of 708 L/ min and a flow velocity of 337 m/s at the enclosure opening. The electrocautery pencil enclosure was also compared to the surgical site evacuators using the same settings.

**Figure 4.**
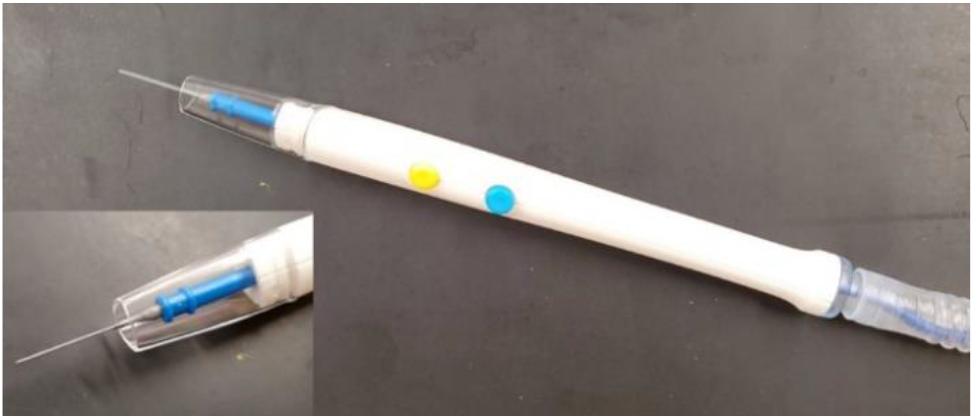
Electrocautery pencil with evacuator port and suction connection.

Skinless chicken breast was used to simulate tissue for electrocautery smoke generation (figure 5). A series of electrocautery power and frequency mode settings were tested. The blend setting corresponded to a 25%-on duty cycle.

**Figure 5.**
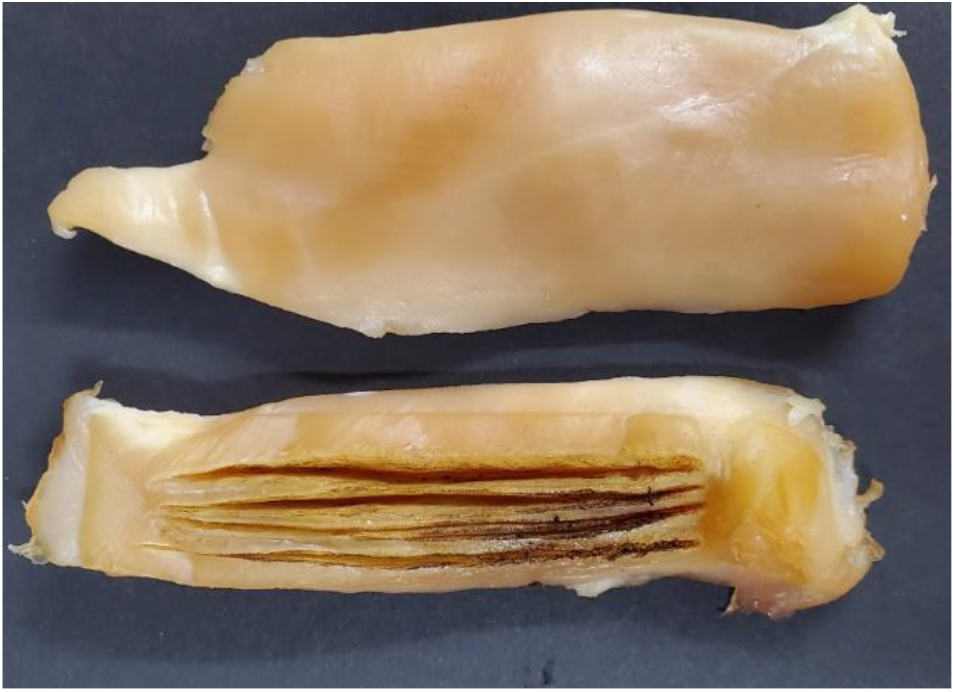
Electrocautery tissue simulation.

## Results

Figures 6 through 12 show frame captured images from corresponding videos that are available in the online supplemental material. The videos are grouped to illustrate the effects of evacuator parameters on aerosol and smoke clearance. Evacuator effectiveness was rated visually based on the presence of particles traversing the field above the upper plane of the evacuator port opening. Complete evacuation indicates that all particles exiting the stoma or tissue entered the evacuator port; no effect implies that the port did not appear to alter the trajectory of the particle stream after it entered the field. Partial evacuation indicates that at least some particles escaped evacuation above the field.

**Figure 6.**
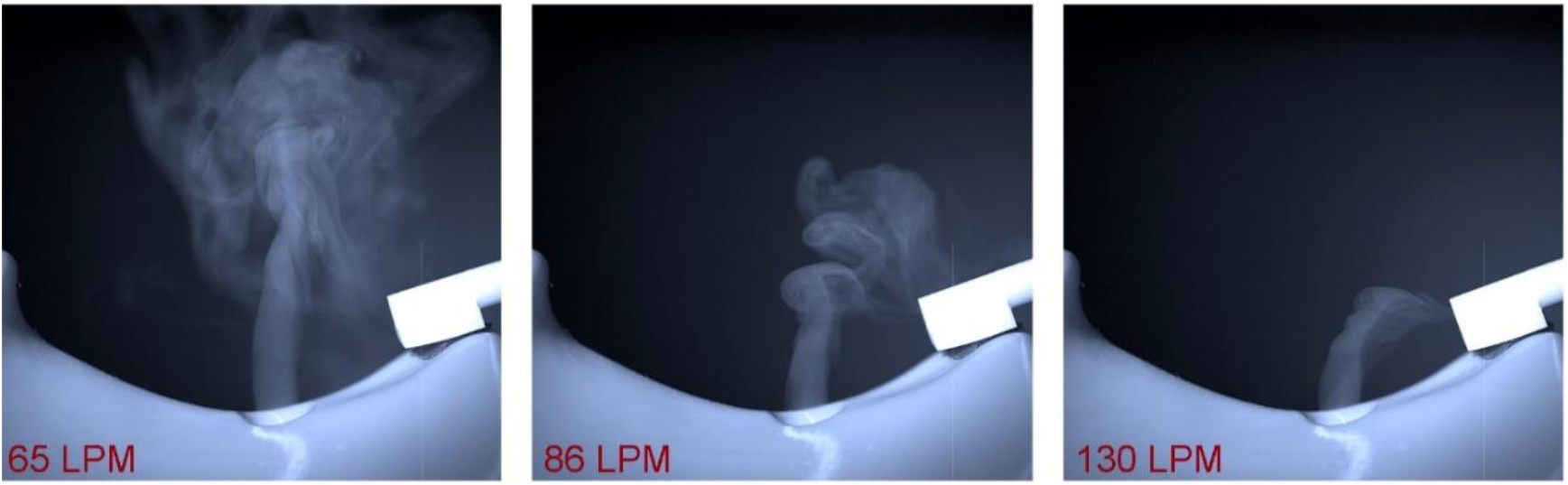
Effect of site evacuator flow rate. Supplement video - Evacuator Flow Rate Effects.wmv.

**Figure 7.**
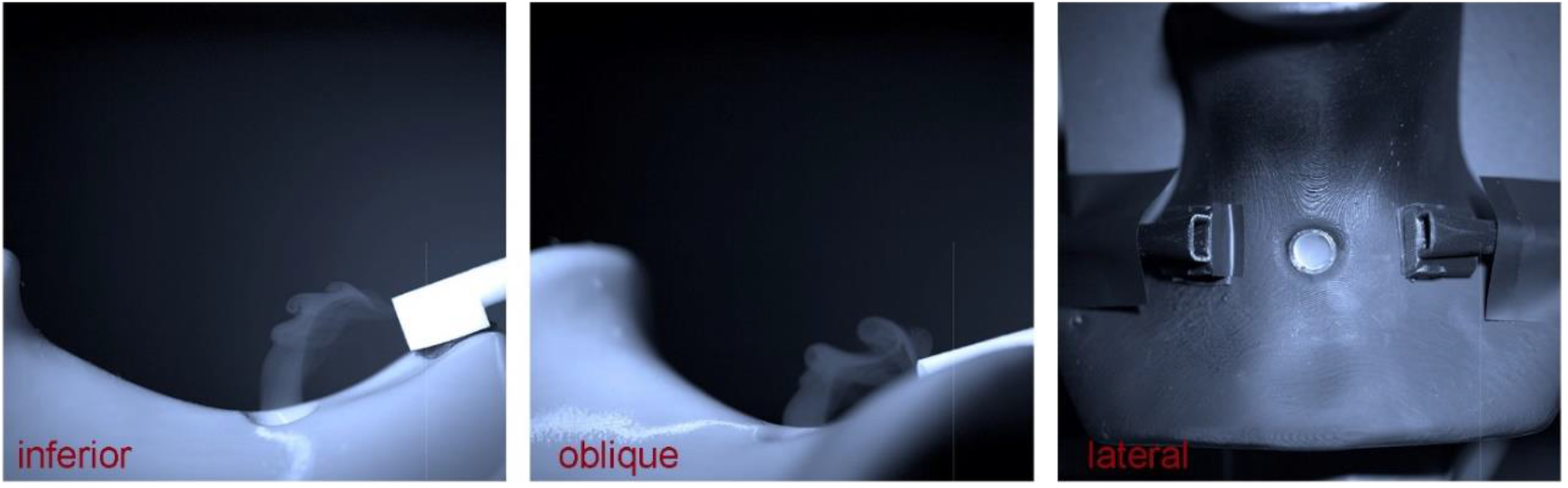
Effect of site evacuator positioning. Supplement video - Evacuator Position Effects.wmv.

**Figure 8.**
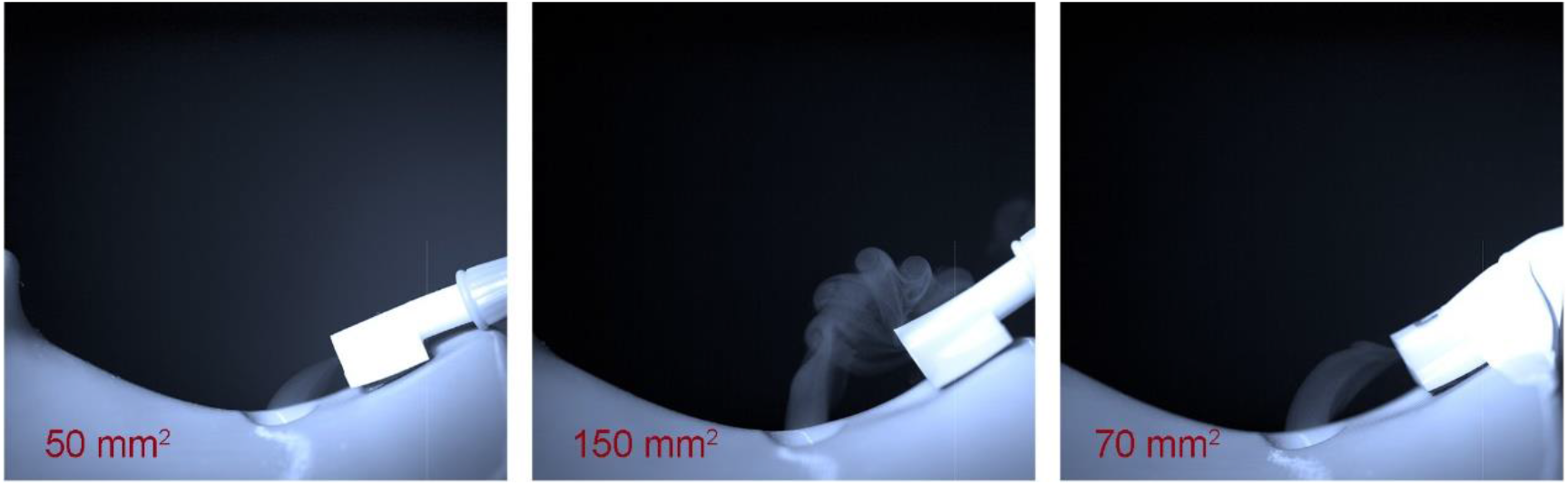
Effect of site evacuator port size. Supplement video - Evacuator Port Size Effects.wmv.

**Figure 9.**
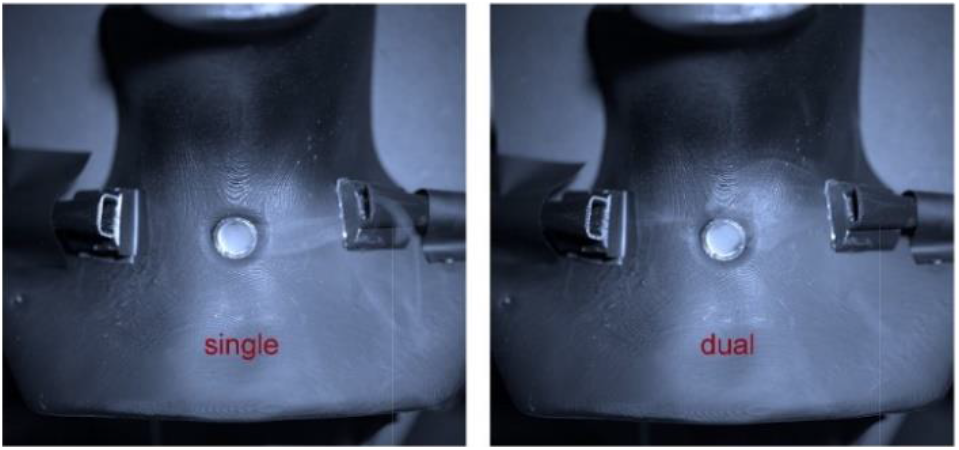
Effect of single vs dual site evacuator. Supplement video - Evacuator Single vs Dual Port Effects.wmv.

**Figure 10.**
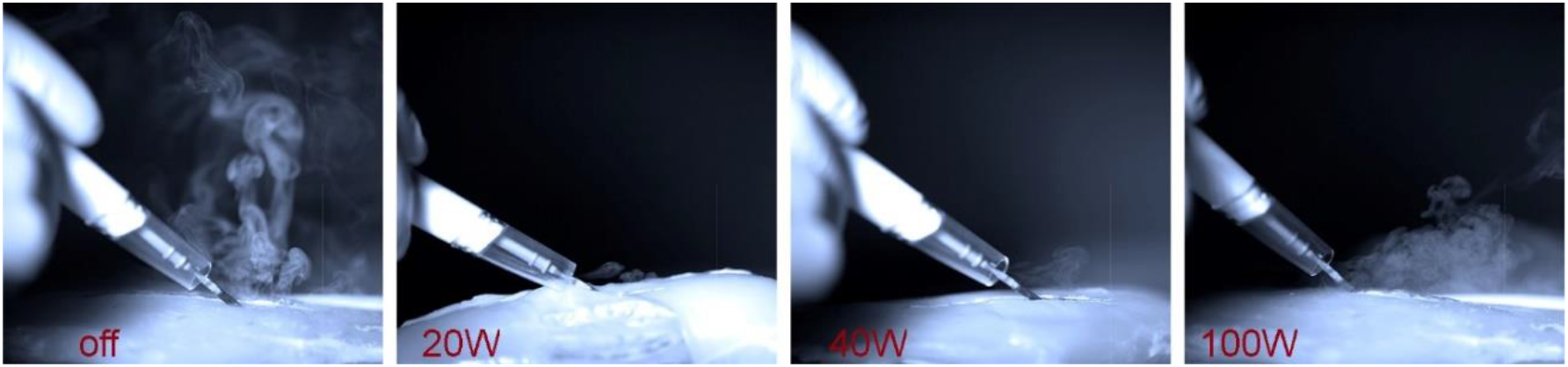
Effect of electrocautery power setting. Supplement video - Electrocautery Power Effects.wmv.

**Figure 11.**
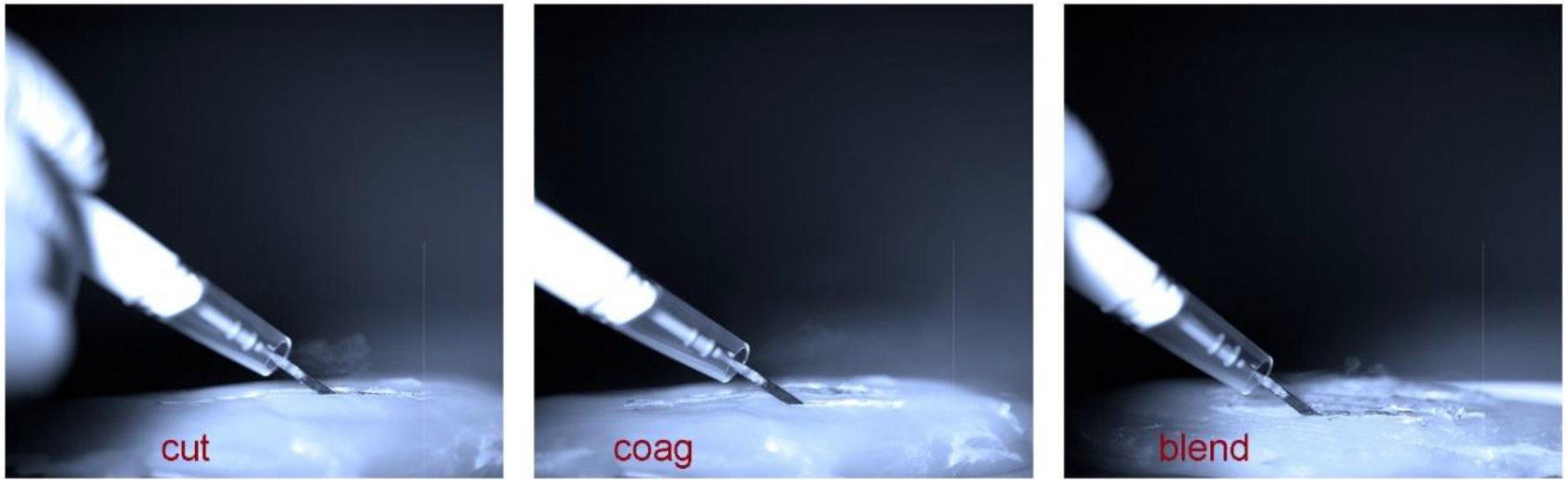
Effect of electrocautery mode (frequency). Supplement video - Electrocautery Mode Effects.

**Figure 12.**
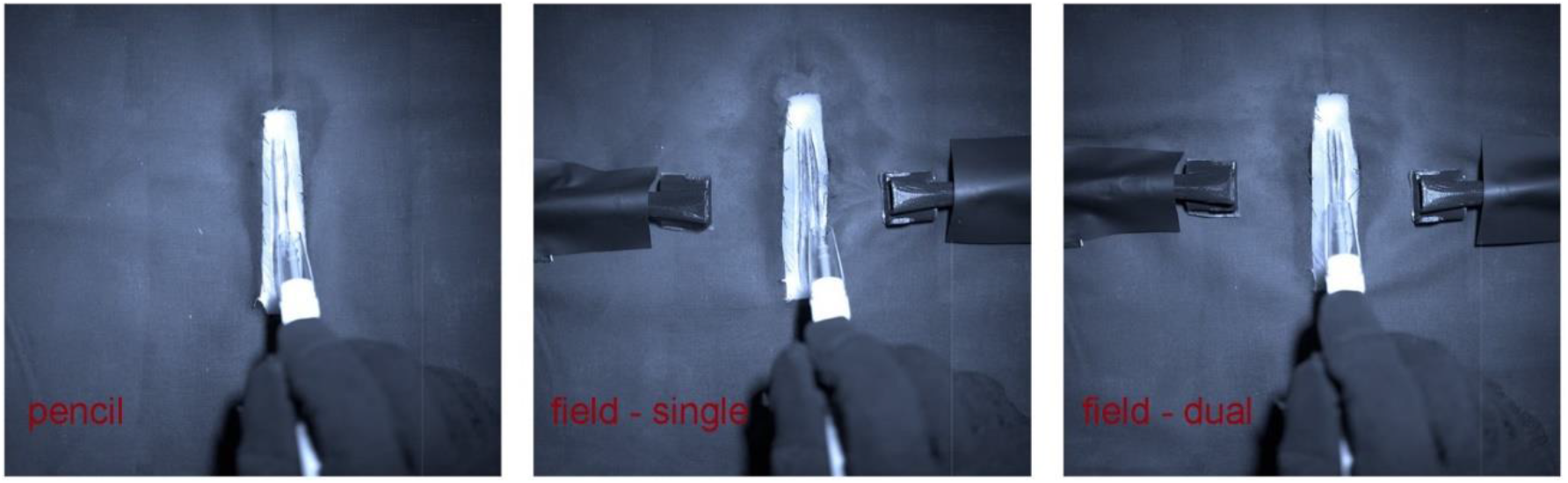
Pencil vs. Field Evacuation. Supplement video - Pencil vs Field Evacuator Effects.wmv

### Surgical site evacuation during aerosol generating procedures (Table 1)

The optimal distance from the opening of the evacuator port to the center of the stoma was 3 cm. In the case of cervical anatomy, inferior placement performed better than lateral or oblique placement. This reflects the elevation of the port relative to the surgical field. Ports with smaller openings performed better than those with larger openings, although the 70 mm^2^ rectangular port appeared to clear the field as well as the 50 mm^2^ circular port. Doubling the number of ports resulted in poorer performance at the 130 L/min setting. However, the dual port system performed well at 708 L/min. These findings reflect the importance of maintaining an adequate flow velocity at the port opening.

**Table 1.** Effectiveness of aerosol evacuation.

| Study | Device (fig. 3) | Parameter | Effectiveness |
| --- | --- | --- | --- |
| *Evac flow* | Site evac C | Flow rate – 65 LPM | Partial |
| *Evac flow* | Site evac C | Flow rate – 86 LPM | Partial |
| *Evac flow* | Site evac C | Flow rate – 130 LPM | Complete |
| *Evac position* | Site evac C | Position - inferior, Flow rate 708 LPM | Complete |
| *Evac position* | Site evac C | Position - oblique, Flow rate 708 LPM | Partial |
| *Evac position* | Site evac C | Position - lateral, Flow rate 708 LPM | Partial |
| *Evac port size* | Site evac A | Port size – 50mm<sup>2</sup>, Flow rate 708 LPM | Complete |
| *Evac port size* | Site evac B | Port size – 150mm<sup>2</sup>, Flow rate 708 LPM | Partial |
| *Evac port size* | Site evac C | Port size – 70mm<sup>2</sup>, Flow rate 708 LPM | Complete |
| *Evac single vs dual* | Site evac C | Port number – single, Flow rate 130 LPM | Complete |
| *Evac single vs dual* | Site evac C | Port number – dual, Flow rate 130 LPM | Partial |
| *Evac single vs dual* | Site evac C | Port number – single, Flow rate 708 LPM | Complete |
| *Evac single vs dual* | Site evac C | Port number – dual, Flow rate 708 LPM | Complete |

In summary, the 70 mm^2^ rectangular port placed approximately 3 cm from the source at a height of approximately 1 cm above the base of the surgical field appeared to achieve the best site evacuation. Increasing the size of the port or doubling the number of ports provides a broader area of field coverage but requires an increase in volumetric flow from the vacuum evacuator to maintain flow velocity.Placing the port below the plane of the source does not clear the field.

### Smoke evacuation during tissue electrocautery (Table 2)

The electrosurgical pencil enclosure achieved complete evacuation at 20W and 40W power settings, and partial clearance at 100W. Cutting mode produces sightly greater plume volume than coagulation or blend modes but smoke evacuation appeared the same for all modes. For surgical smoke evacuation, the field evacuator appears to perform as well as the pencil enclosure.

**Table 2.** Effectiveness of electrocautery smoke evacuation.

| Study | Device | Parameter | Effectiveness |
| --- | --- | --- | --- |
| *Electrocaut power* | Pencil evac | Power – 0W | No effect |
| *Electrocaut power* | Pencil evac | Power – 20W, cut | Complete |
| *Electrocaut power* | Pencil evac | Power – 40W, cut | Complete |
| *Electrocaut power* | Pencil evac | Power – 100W, cut | Partial |
| *Electrocaut mode* | Pencil evac | Mode – cut, 40W | Complete |
| *Electrocaut mode* | Pencil evac | Mode - coagulate, 40W | Complete |
| *Electrocaut mode* | Pencil evac | Mode - blend, 40W | Complete |
| *Electrocaut* | Pencil evac | Device – pencil evac, 40W, cut | Complete |
| *evac device* |  |  |  |
| *Electrocaut*<br>*evac device* | Site evac C | Device – single site evac, 40W, cut | Complete |
| *Electrocaut*<br>*evac device* | Site evac C | Device – dual site evac, 40W, cut | Complete |

## Discussion

For clinical purposes, an aerosol is defined as an airborne cloud of particles suspended in liquid droplets expelled from the respiratory tract. The droplet suspended particles typically consist of materials that are in the respiratory tract including epithelial cells, carbohydrates, proteins, lipids, and microorganisms. Human disease can be spread by bio-pathogens suspended in the droplets. Viral particles range in size from .02 to .2 micrometers. Bacteria range from about 1 to 10 micrometers. The aerosolized droplets are spherical and typically measure 5 to 500 micrometers in diameter. Hence, prior to evaporation, droplets are substantially larger than the pathogens they carry and can be visualized using HSP.

Three primary models of respiratory pathogen transmission by droplets have been identified^8^. *Contact transmission* refers to physical contact with respiratory fomites (e.g., hand to mouth or nose). *Direct droplet transmission* occurs when pathogen containing liquid droplets are deposited onto respiratory mucus membranes. *Airborne transmission* refers to inhalation of air that contains suspended droplet nuclei. For a given pathogen, the transmission modes are not mutually exclusive and spread varies with the local environment in the region of the source.

Smoke generated in the surgical field also has the potential to carry bio-pathogens into the local surgical environment around the patient. Smoke differs from respiratory aerosols in that it is the result of tissue vaporization from heat energy. The resulting plume can carry viral particles via convection. Particles are not suspended in liquid droplets and are not propelled by mechanical forces. Additionally, smoke plume contains potentially harmful hydrocarbons that can contaminate the local environment via diffusion.

There are important differences between aerosols and smoke in the operating room. Pathogen laden aerosol droplets can be trapped by surgical personal protective equipment while potentially harmful substances in surgical plume may escape filtration. The significance of hydrocarbons produced from tissue vaporization during surgery is unclear, but these volatile substances are probably not cleared by currently available environmental measures to protect healthcare workers during surgery.

Studies of static aerosol distribution have used droplet collection techniques (glass slides, enclosure walls) to estimate the size and number of droplets in the region of an aerosol source^9^. Aerosol mass spectroscopy^10^ is a sensitive measure of the composition and quantity of particles suspended in air that is commonly used in environmental air quality studies. The most used device, the optical particle counter (OPC), is based on particulate scattering of incident light in an enclosed chamber^11^. OPC uses a photoelectric sensor that detects the light scattering. The device converts the light reception into a voltage signal that varies directly with the particle size. Except for mass spectroscopy, these aerosol detection methods are relatively simple and inexpensive to implement. However, they also do not provide any information regarding particle movement over time relative to the aerosol source.

### High speed photographic analysis of bioaerosols

Methods to visualize aerosol production include high speed photography (HSP)^12,13,14,15^, light-sheet analysis within a confined chamber^16,17^ or under a microscope^13^, and aerosolization of fluorescein solutions^18^. For direct visualization of particle dynamics, high-resolution HSP is required. In addition to particle distribution dynamics, HSP has been used to measure particle velocities (particle image velocimetry). Evaluating the effects of surgical evacuation differs from similar studies of aerosol distribution during normally occurring respiratory events (breathing, coughing, sneezing). Most quantitative studies of respiratory droplets focus on the epidemiology of disease transmission^19^ where the distribution of potential pathogen bearing particles in the surgical field over time may be more relevant to understanding intraoperative transmission risk.

Studies of naturally occurring respiratory events are typically concerned with the distribution of particles as a function of distance from the source, whereas evacuation of the surgical site is intended to clear the immediate vicinity of the wound. In the case of simulated AGPs, our study documents the effectiveness of droplet clearance within 1 cm from the plane of the surgical wound. Clearance is achieved before the potential droplet cloud can be inhaled by surgical team members, assuming their nose/mouth is approximately 30 cm from the field.

### Protecting health care workers

The primary means of protecting health care workers in the operating room from contaminated aerosol and smoke is personal protective equipment. For high risk AGPs, high filtration respirators and powered air purifying respirators are recommended. However, these can increase the user’s work of breathing and can lead to operator discomfort, distraction, and fatigue during longer surgical procedures. They can be intrusive and introduce technical surgical limitations, especially during microscopic, endoscopic, and other minimally invasive procedures. These devices are also subject to cost and availability issues.

The other means of protection for operating room personnel is negative pressure ventilation^20^. This is typically designed with other environmental measures such as equipment isolation and limited OR traffic. The effectiveness of negative pressure environments in protecting health care workers has not been documented and there is the possibility that the surgical team is in the path of the bioaerosol flow. Most surgical suites are not equipped with negative pressure ventilation capabilities and retrofitting can be complex and expensive.

### Future studies

Our research group is currently working on quantitative image processing algorithms to measure particle clearance and optimize the evacuator designs. Particle velocimetry will enable us to determine the extent of flow directional change in the region of the evacuator opening. Using the technical knowledge gained from these experiments, we also plan to complete similar studies of other surgical fields. We have designed prototypes for trans-nasal and oral cavity procedures (figure 13). The designs are suitable for 3D printing and prototyping. HSP techniques learned during this study will allow us to study the production and clearance of aerosols during other AGPs.

**Figure 13.**
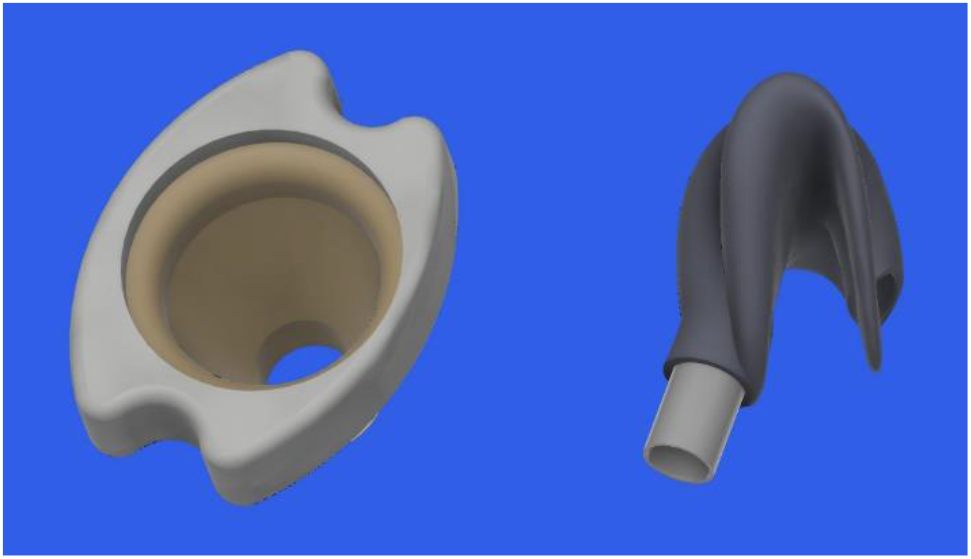
Nasopharyngeal and oral cavity surgical site evacuator designs.

## Conclusions

High speed photographic imaging of the surgical site documents the potential for evacuation of aerosols and plumes produced by aerosol generating procedures and smoke producing surgical devices. Effective surgical site aerosol evacuation requires careful design of the evacuator ports and proper placement in the field. The commercially available evacuation enclosure used in this study is effective for electrocautery smoke clearance.

## Data Availability

Data relevant to this study will be made available online once the manuscript is published.

## Acknowledgements

Symmetry Surgical Corporation – supplied smoke evacuation, electrocautery, and high-speed photography equipment.

Louisiana State University Health Sciences Center– resident research grant.

